# A Double Power Law model accurately forecasts COVID-19 transmission and mortality

**DOI:** 10.1101/2020.05.07.20094714

**Authors:** Vladimir A. Osherovich, Joseph Fainberg, Lev Z. Osherovich

## Abstract

The novel coronavirus SARS-CoV-2 appeared at the end of 2019, spreading rapidly and causing a severe respiratory syndrome (COVID-19) with high mortality. Until a vaccine or therapy is found, the most effective method of prophylaxis has been to minimize transmission through mandatory social distancing and restriction of all but essential economic activity. A key question facing policy makers and individuals is when to resume economic and social activity in the face of persistent community transmission of SARS-CoV-2. To help address this question, we have developed a mathematical model of transmission and mortality of COVID-19 in countries that have implemented stringent social distancing measures. Using data from Italy, Spain, Switzerland and Germany on SARS-CoV-2 transmission, active caseload and mortality, we model the rapid rise and slow decay (“long tail”) of the COVID-19 pandemic using a first order nonlinear differential equation. The prognostic utility of our model is validated by strong correspondence between predicted and prospectively observed data up to eight weeks after curve fitting. This Double Power Law model can be applied to other countries as a predictive tool to inform policy decisions concerning social distancing.

## 1. Introduction

Social distancing measures, implemented at different times and to varying degrees world-wide, have reduced transmission of SARS-CoV-2 compared with early phases of the pandemic, resulting in “flattening of the curve” of new infections and gradual reduction in mortality. The observed long (3-5 day) asymptomatic incubation period and the variable (5-14 day) period of progression from exposure to death make it difficult to model the individual biological and behavioral components that influence SARS-CoV-2 transmission. Because of the very large number of cases and generally high accuracy of publicly available case data, it is instead possible to model the COVID-19 pandemic as a multivariate statistical phenomenon without explicitly defining and parameterizing the numerous factors influencing transmission and mortality.

We have analyzed emerging data from European countries that initially experienced uncontrolled community transmission and subsequently implemented rigorous social distancing measures and nearly complete shutdown of nonessential economic activity. At the time of our analysis (mid-April 2020), the two European countries which had experienced the earliest and most severe outbreaks of COVID-19 were Italy and Spain, which respectively reported their first cases on February 22nd and 25^th^ and reached peak case load on March 21st and March 26th. By mid-June, both countries reported low levels of infections (<5% daily new cases compared with peak) and had substantially eased their social distancing measures, thus providing a comprehensive data set with which to model the initial phases of SARS-CoV-2 and the effect of social distancing measures.

To model the infection rate and death rate curves for Italy and Spain, we treat the spread of SARS-CoV-2 as a multiscale process with two time scales: one time constant (m1) defines the growth rate and another time constant (m3) defines the relaxation process that results from effective social distancing measures; m1 is on the order of a few days while m3 is at least one order of magnitude larger (30-60 days). The other two constants (m2 and m4) are non-dimensional and relate to the powers of growth and relaxation phases. This formula, termed the Double Power Law (DPL), is given as (1):

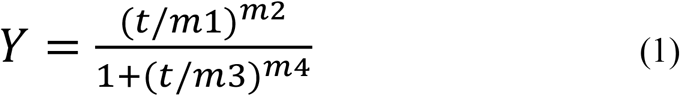

where the evolution function Y can be for new cases per day (NC), daily death rate (DR) or active cases (AC). The DPL Formula (1) is an exact analytical solution for the special case of the first order nonlinear ordinary differential equation named after Jacob Bernoulli (Bernoulli, 1695):

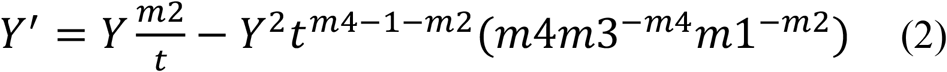

In Formula (2), the left side is the time derivative of Y. On the right side a linear term is responsible for the initial growth and a nonlinear negative term controls the relaxation. Both terms (linear and quadratic) contain some powers of time t. The general Bernoulli equation contains two arbitrary functions of time and the nonlinearity does not have to be quadratic for Y.

## 3. Results

To obtain parameters for the DPL formula, we analyzed data from Italy and Spain, two countries that experienced rapid initial growth of NC and instituted rigorous social distancing early during the pandemic. Importantly, the primary data span times before and after NC and DR peaks, thus providing a most complete picture of COVID-19 dynamics before and during social distancing measures.

Figures 1a and 1b respectively show the NC and DR for Italy. The black circles are daily values averaged over 3 days for the period from February 22, 2020 through April 19, 2020, which was sufficient to define the parameters m1, m2, m3 and m4 (Table 1). For both NC and DR, the resulting curve was extrapolated (in red), showing a high degree of correspondence with prospectively gathered data for up to two months. The regression coefficient (R) for the NC fit in Figure 1a is R=0.98. DR observations (Figure 1 b black circles) also show strong regression (R=0.99) for data gathered after April 19. The projection of DR out to 100 days after February 22, 2020 is asymmetric and shows a long tail, consistent with observed persistence of community-acquired SARS-CoV-2 infection. Overall, the projected number of total deaths (TD) at 100 days is 3.68 times larger than the total deaths at the peak of DR (Figure 1c and Table 3).

**Table 1.** Fit Parameters for New Cases and Death Rates.

| Data | Italy | Italy | Spain | Spain |
| --- | --- | --- | --- | --- |
|  | NC | DR | NC | DR |
| m1 | 2.49 day | 2.21 day | 2.79 day | 3.76 day |
| m2 | 3.83 | 2.97 | 4.29 | 4.11 |
| m3 | 27.59 day | 26.0 day | 26.1 day | 22.86 day |
| m4 | 5.45 | 4.58 | 6.35 | 6.15 |
| R | 0.98 | 0.99 | 0.98 | 0.99 |
| sigma | 370 per day | 42 per day | 562 per day | 30 per day |
| t<sub>max</sub> | 32.3 day | 29.25 day | 29.1 day | 25.61 day |
| Y<sub>max</sub> | 5463/day | 778/day | 7723/day | 877/day |

**Table 2.** Fit Characteristics for New Cases (2020)

| Data Fit | Italy Feb 22 to Apr 19 | Spain Mar 1 to Apr 19 |
| --- | --- | --- |
| Peak day | March 24 | March 29 |
| Peak value | 5455 per day | 7722 per day |

**Table 3.** Fit Characteristics for Death Rates and Total Deaths (2020)

| Data Fit | Italy Feb 28 to Apr 19 | Spain Mar 6 to Apr 19 |
| --- | --- | --- |
| Peak day | March 28 | March 31 |
| Peak value | 777 deaths/day | 871 deaths/day |
| Sum: Day 0 to Peak day | 9722 deaths up to Peak | 8417 deaths up to Peak |
| Sum: Peak Day to day 100 | 27082 deaths after Peak | 22646 deaths after Peak |

**Figure 1.**
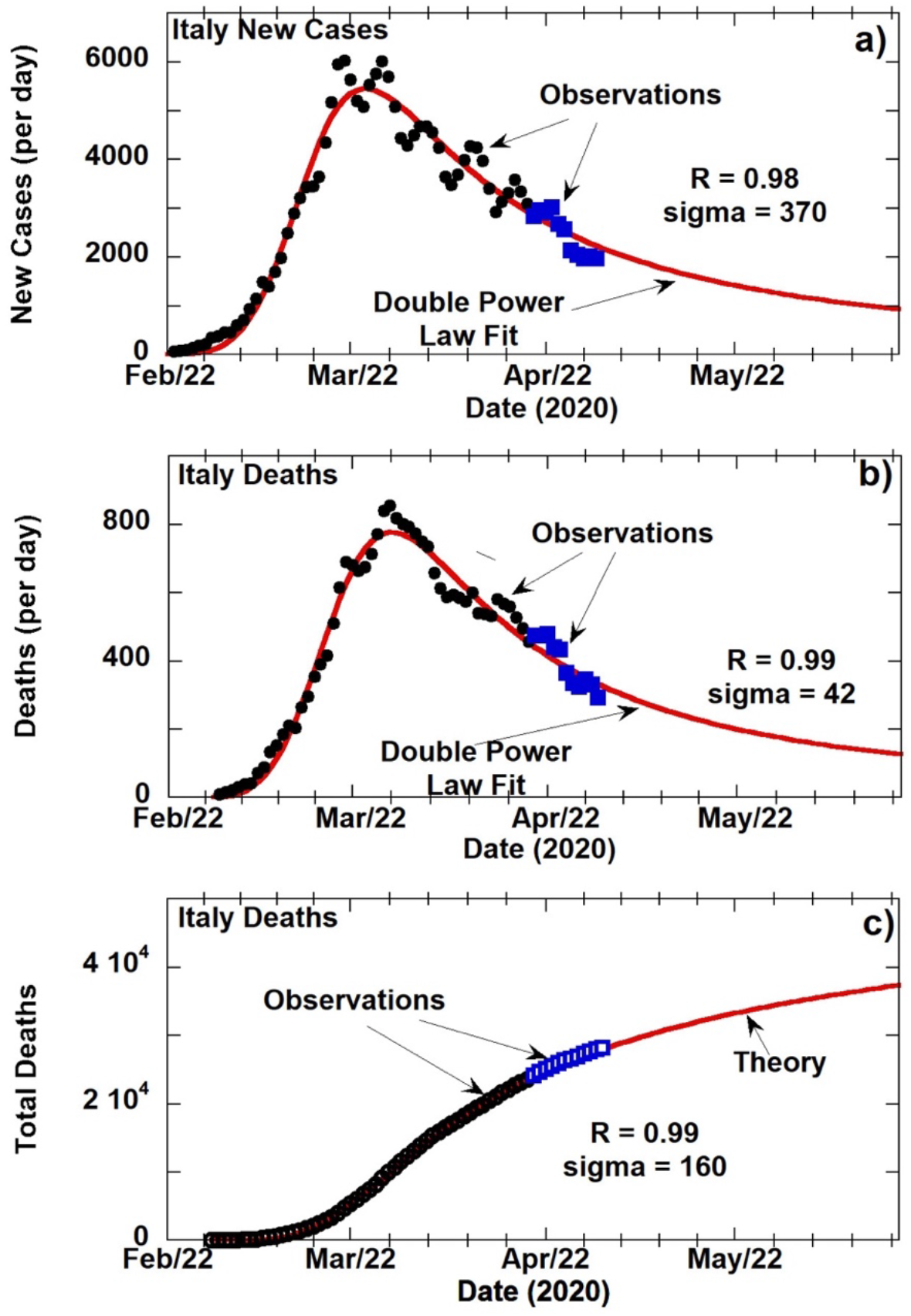
**a)** COVID-19 new cases per day in Italy with 3-day averaging starting from February 22, 2020; **b)** Death rate (DR), defined as deaths per day, in Italy starting at February 28, 2020; **c)** Cumulative death numbers from February 28, 2020 in Italy. Black circles in a) and b) are the observed values up to April 19, 2020 that were used for the DPL curve fitting (red lines). Blue squares are values observed after April 19, 2020 and were not used in fitting. In c) open circles represent total death numbers summed up without 3-day averaging for that date with blue squares for data after April 19, 2020. In Figures 1a and 1b, the red line is the resulting theoretical fit of the 3-day average daily NC and DR data for Italy. In Figures 1a and 1b, red lines depict fitting of the sampling interval February 22, 2020 to April 19, 2020 extrapolated into the future. Starting point for NC and the DR is at February 22, 2020 and February 29, 2020 respectively. The horizontal scale corresponds to a 100-day evolution of both NC and DR. Data obtained after construction of the model are shown as blue squares (labeled as “Observations”) and follow closely the theoretically extrapolated curve. The black circles and blue squares are 3-day averages of the observed and projected values, respectively. The solid red curve is the projection of the DPL model beyond April 19. For each curve, t_tmax_ is the time of peak (Y_max_). Values for t_tmax_ are given in Table 1. Peak dates and new case load at t_tmax_ are shown in Table 2. In Figure 1c, the total (cumulative) number of deaths (TD) is shown by the red line which has been extended out to 100 days from the beginning. Black circles are the sum of reported daily deaths up to that date. Root mean square deviation of observed data is given as sigma on the plots and in Table 1.

The results for Spain (Figure 2) are similar to those of Italy for both DR and TD. Figure 2b shows a somewhat faster growth of DR at the beginning (March 1, 2020); m2 obtained for Italy is smaller compared to m2 for Spain. The relaxation for Spain appears shorter since its value for m4-m2 is slightly larger than the value obtained for Italy. The projection of the ratio of total deaths at day 100 to the total deaths at the day of the DR peak for Spain is 3.69, slightly larger than for Italy. The sudden surge of new cases (bump on tail) after April 19 is shown in Supplementary Data 2 together with weekly oscillations observed in 3-day averaged data.

**Figure 2.**
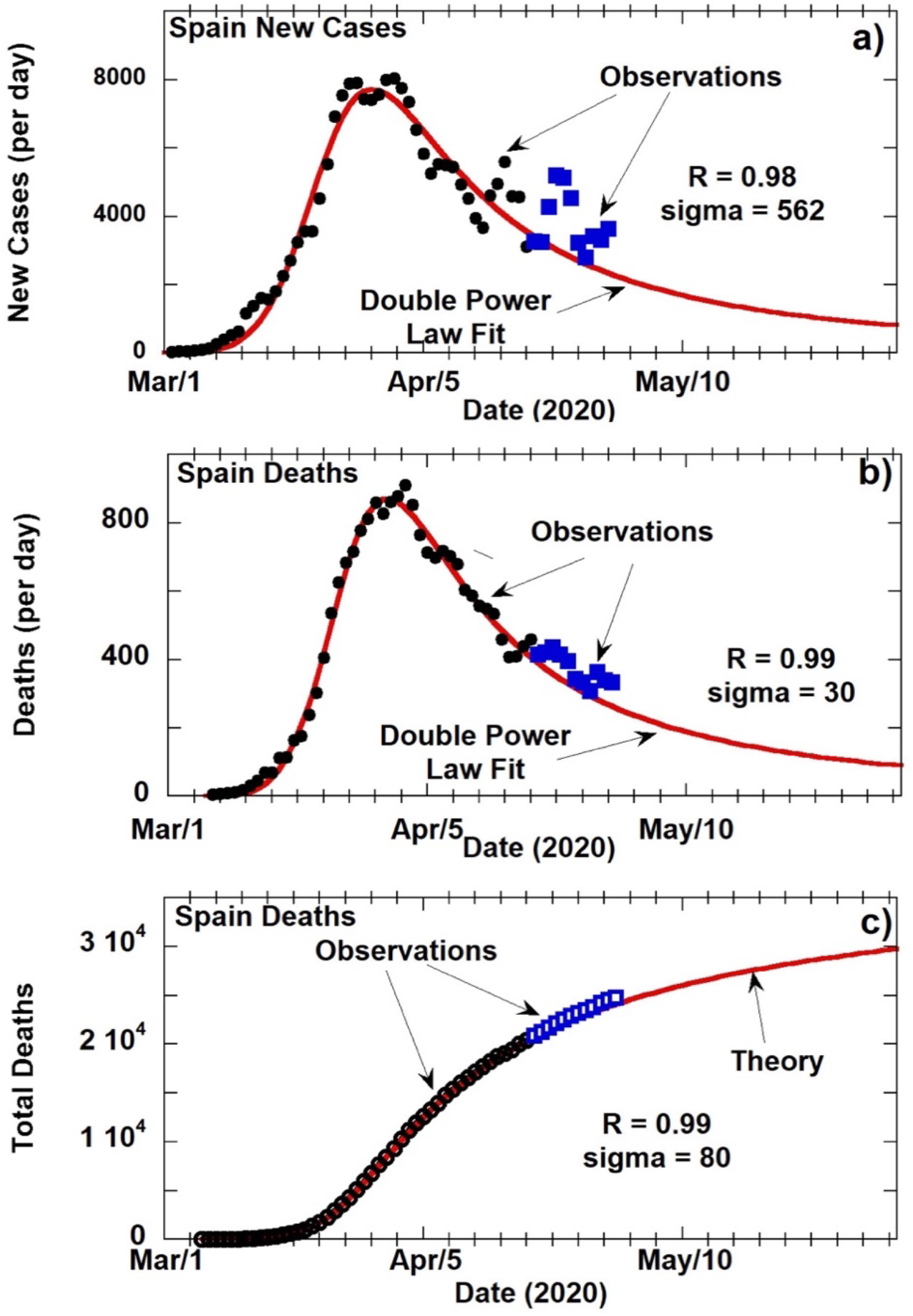
**a)** COVID-19 new cases per day in Spain with 3-day averaging starting from March 1, 2020. The blue squares represent data after April 19, 2020 and represent the large 7-day oscillation for Spain seen in Figure 3; **b)** Death rate (deaths per day) in Spain; **c)** Total death number from March 6, 2020 in Spain. Legends and methods are same as in Figure 1.

An additional metric of COVID-19 prevalence is Active Cases (AC), defined as total cases - (deaths + recovered cases). Using retrospective AC data from Italy, Spain, Germany and Switzerland, we find that AC also follows DPL behavior (Figure 4). Black circles correspond to observations and red lines are the corresponding DPL fits. Table 4 gives values for m1, m2, m3 and m4 for the DPL fits for AC in all 4 countries.

**Table 4.**
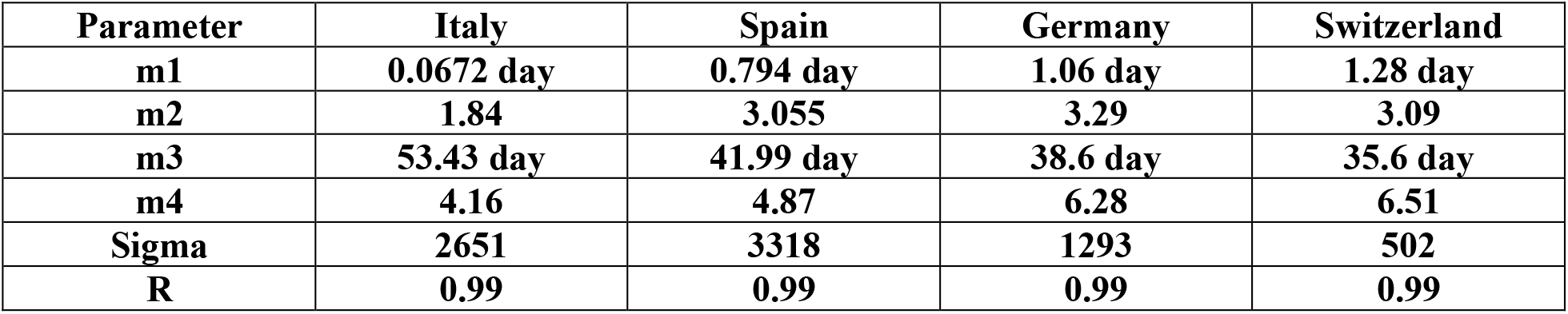
Active Cases (number of infected people)

**Figure 3.**
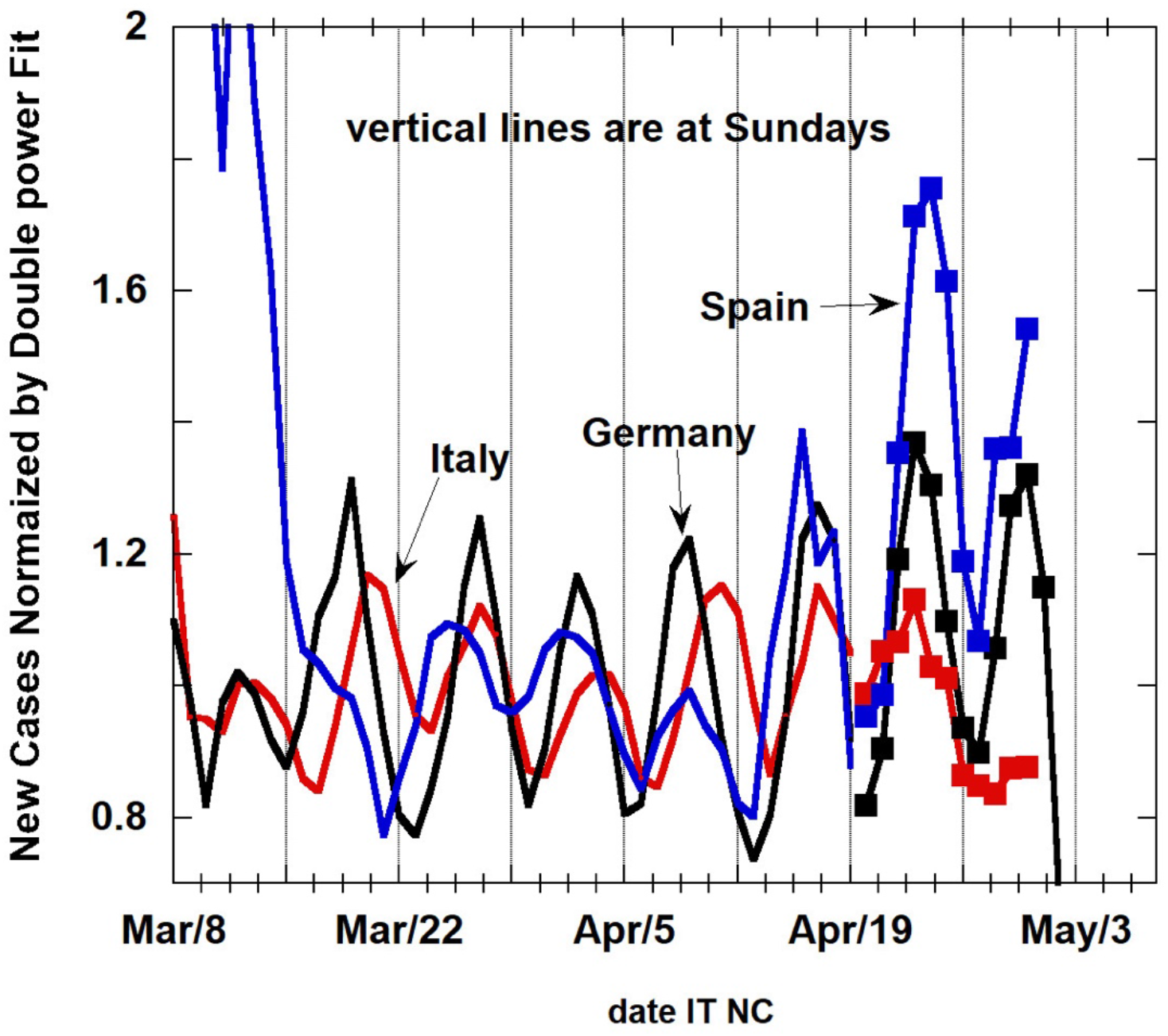
Weekly oscillations of new cases in Italy (black), in Spain (blue) and in Germany (red) with Sundays shown by vertical lines. The 3-day averages are normalized by the Double Power Law fit values. The squares are observed values after April 19, 2020 which was the end dates for the fits. Spain shows large weekly oscillations in late April. The minima of these oscillations appear near weekends.

**Figure 4.**
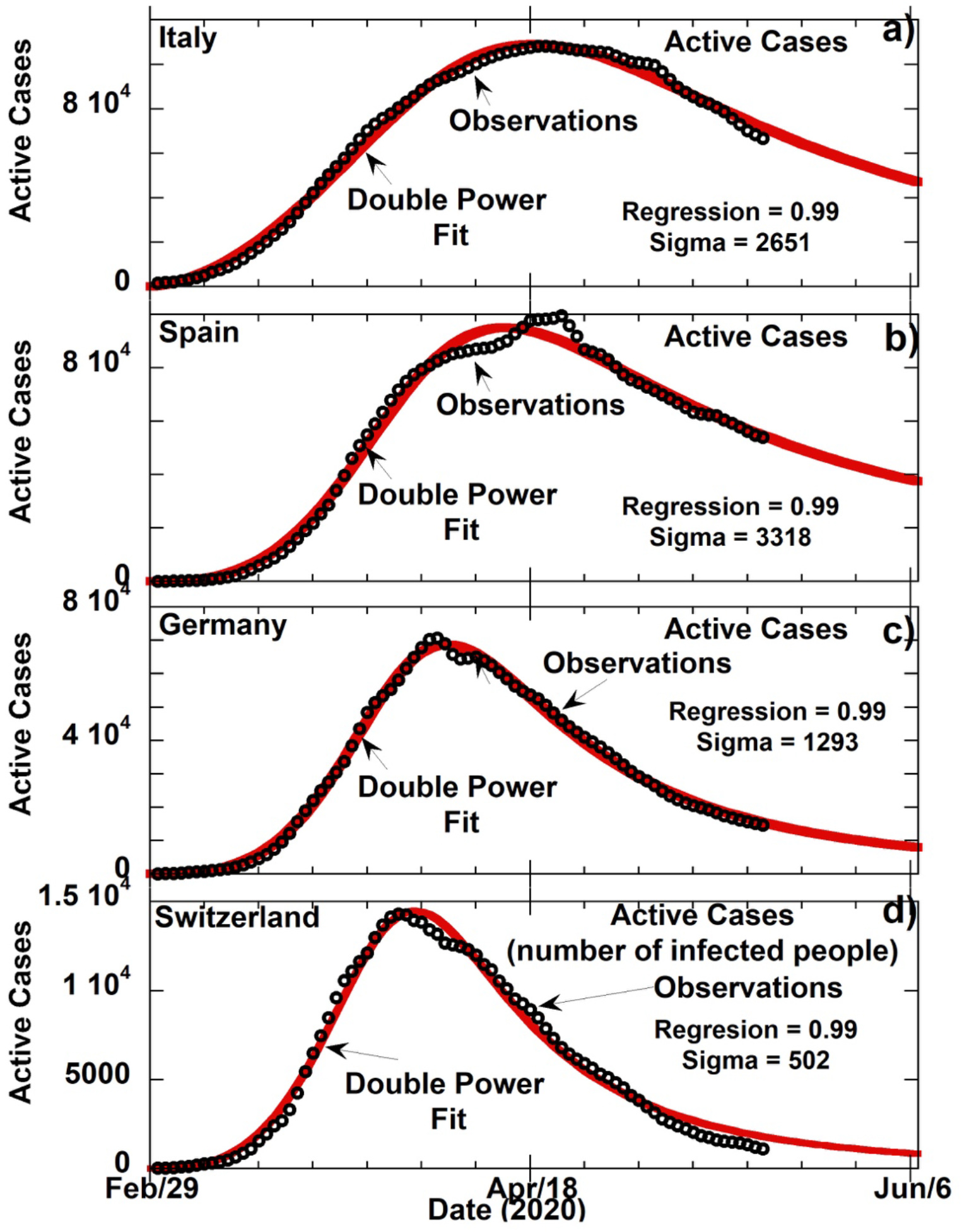
COVID-19 Active Cases (number of infected people) for Italy, Spain, Germany and Switzerland. Active Cases are defined as total cases – total deaths – recovered cases. Black circles are 3-day averages of observations. Red lines are DPL formula curve fits extrapolated past the April 19 curve fitting cutoff. Corresponding fit parameters m1, m2, m3 and m4 are in Table 4.

## 4. Discussion

In this paper we have suggested an analytic formula (1) that describes the epidemiological evolution of COVID-19 for a given country, accurately forecasting new cases or death rates (Y). Often the early phase of growth is described as exponential, but our curve fitting shows that growth of COVID-19 more accurately follows a power law, as does the decrease of Y after the peak time. At the beginning of the process (small t and m2>0), prior to the implementation of social distancing measures, the model follows an asymptotic power law ∼ (t/m1)^m2^ that governs the growth phase. When effective social distancing measures are in place (large t and m2-m4<0), Y decreases according to a second power that governs relaxation phase, as *Y*∼*t*^*m*2−*m*4^*m*3^*m*4^*m*1^−*m*2^ per the Bernoulli equation. Because of the concurrent effect on Y from separate power laws for growth and relaxation, we refer to Formula (1) as a Double Power Law.

The DPL model has high predictive value – for example, at time of submission of this manuscript and 8 weeks after curve fitting, the DPL projections for TD in Italy and Spain are accurate to within 10% of observed data (Italy TD (observed)=34301 vs TD (projected)=36873 and Spain TD (observed)=27136 vs. TD(projected)=30191). The high degree of correspondence of prospective data (blue squares) to the projections of DPL curves for NC and DR validates the DPL model and allows extrapolation into the future, providing that social distancing measures remain in place. For NC and DR of both Spain and Italy, we observe that m3 is comparable to t_max_, suggesting that social distancing measures and other factors governing relaxation were similarly effective in these two countries.

Given the high level of prognostic accuracy of the DPL model, these projections can serve as a baseline to model how withdrawal of social distancing measures will impact new cases and deaths. We hypothesize that regional variations in implementation and adherence to social distancing measures will impact parameter m3 and m4 (related to relaxation) rather than m1 and m2 (related to growth). Additionally, we have noticed the phenomena of weekly oscillations in NC and DR for Italy, Spain and Germany when. 3-day averaging is used (Supplementary data 2). The nature of these oscillations will be addressed in future work (manuscript in preparation).

Other researchers have attempted to model dynamics of the SARS-CoV-2 pandemic, focusing on theoretical and practical factors governing transmission rates. For example, social distancing as a means of reducing SARS-CoV-2 transmission has been modelled by Ferguson *et al*. 2020, Flaxman *et al*. 2020 and Hsiang *et al*. 2020. Kissler *et al*. 2020 model the effects of seasonal transmission, acquired immunity and non-pharmaceutical interventions such as social distancing on viral reproductive number R_0_. We are not aware of previous efforts to apply Formula (1) as the solution of the Bernoulli differential equation for modeling of COVID-19 or any other epidemiological processes.

Critiques of the use of power law models in biological phenomena focus on lack of statistical validation for such approaches and lack of underlying mechanistic knowledge about the parameters that drive these models (Stumpf and Porter, 2012). While we acknowledge that complex biological and social factors influence SARS-CoV-2 transmission, we explicitly do not attempt to model these individually, instead treating growth and relaxation as distinct composite processes. Moreover, there is strong statistical validation of the Double Power Law for COVID-19 incidence and mortality. Firstly, we have reproduced the observed death rates and total deaths with high accuracy (R>0.98) for two countries up to April 19 using the DPL formula. Secondly, our model accurately predicts data gathered after April 19, with >90% accuracy at >8 weeks after curve-fitting. We hypothesize that while growth parameters reflect fundamental biological and behavioral factors involved in viral transmission, relaxation parameters reflect the efficacy of social distancing, but more work is needed to define and parametrize the specific biological, behavioral and policy components that govern COVID-19 dynamics.

A key question facing policy makers and individuals is when to resume previous economic and social activity in the face of persistent community transmission of SARS-CoV-2. The practical implications of the DPL model for public policy are two-fold. First, the high predictive accuracy of the DPL model should help policymakers to anticipate the likely range of case load and mortality after an extended period of social distancing. The observed slow decay of case numbers even after stringent social distancing measures reveals limitations to non-pharmaceutical interventions to control transmission. The DPL model predicts that even if social distancing is rigorously maintained, the number of COVID-19 deaths after DR peak may be 2 – 3 times larger than the total number of deaths up to the peak. Second, the DPL model should inform rational decision-making about when to ease social distancing measures. As many countries are now relaxing their social distancing measures, we expect that the DPL model could be used to parametrize and compare the effect of different policies on NC and DR. Premature slackening of social distancing measures is expected to cause resumption of growth phase, resulting in increased cases and deaths, but it remains unclear when there is unlikely to be further benefit to continue with social distancing. The DPL model’s predictive utility diminishes as NC and DR approach the root mean square deviation of observed data, given as Sigma in Table 1. However, if social distancing measures are withdrawn before NC has reached Sigma, the DPL model predicts that growth will resume according to the first power law component, although the impact of resumed growth on NC would not be apparent for at least two weeks. Likewise, when new vaccines and therapies are developed and deployed, changes in the parameters of the DPL model can be used measure the impact of these interventions on NC and DR. We intend to apply the methodology developed in our paper to study COVID-19 dynamics in other countries like the United States and Brazil which have struggled to effectively implement social distancing (manuscript in preparation).

## 5. Methods

Caseload and death data is drawn from Johns Hopkins University Coronavirus Resource Center (https://https://coronavirus.jhu.edu/map.html) and https://www.worldometers.info/coronavirus/.

## Data Availability

Data is drawn from Johns Hopkins Coronavirus Resource Center and compiled on https://www.worldometers.info/coronavirus/

## 6. Supplementary Data 1

The maximum of NC and DR according to Formula (1) is described as

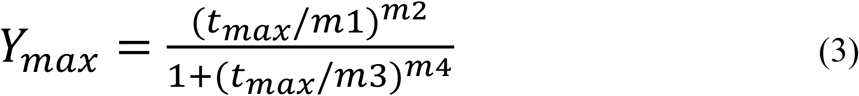

where t_max_ is the time when Y_max_ occurs. At the time of the maximum, the first derivative of Y must be zero. The last condition allows us to express t_max_ in terms of the constants m1, m2, m3 and m4

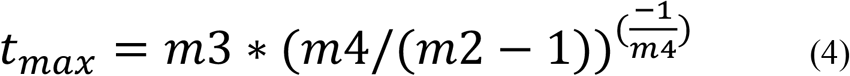

Formula (4) allows us to express m2 as a function of m3, m4 and t_max_. Thus, theoretically instead of m1, m2, m3, and m4 as fitting parameters, the set of 4 numbers Y_max_, t_max_, m1 and m3 can be used. It is important to recognize that once Y_max_ and t_max_ are known, together with the whole growth part of the curve up to the peak, then the relaxation part of the curve is completely determined.

## 7. Supplementary Data 2

For new cases presented in Figure 1a for Italy, observational data points (black circles) form a double peak. In fact, there are 7-day oscillations around the smooth double power fit particularly evident during the relaxation phase of the NC curves. For Death Rate (Figure 1b), these oscillations are also present. In Figure 3, we show NC normalized by the Double Power fit for three cases. The black curve is for Italy, the red is for Germany and the blue is for Spain. We verified that NC for Germany also fits well with the DPL Formula (1). We also observed weekly oscillations in data for the NC and DR for other countries including the United States. Examining this in detail is beyond the scope of this paper. Normalization presented in Figure 3 will be used to account for weekly oscillations even when the amplitude of oscillations is large.

## Acknowledgements

The authors are grateful to Elena Romm from NIH/National Institute of Allergy and Infectious Diseases and Jordan Fainberg for discussions and suggestions.

